# Real-world drug use in ATC and ICD-10: an expert-curated drug–diagnosis resource based on UK primary care and Danish hospitalization electronic health records

**DOI:** 10.64898/2026.09.06.26362128

**Authors:** Ioannis Louloudis, Hannah Currant, Cai R Ytsma, Sidsel Christy Lindgaard, Sedrah Butt Balaganeshan, Amalie Dahl Haue, Maria Pisliakova, Wentin Chen, Samuel Thio, Søren Brunak, Spiros Denaxas

## Abstract

As the use of electronic health records in drug repositioning research increases, so does the need for a well-curated resource describing real-world drug-diagnosis relationships. This need is particularly important in the context of polypharmacy. Although literature-based drug-disease maps exist, they are typically based on mechanistic disease ontologies, which are not widely used in clinical settings and do not align well with the ICD system, which is most often used in healthcare. Here, we used real-world primary and secondary healthcare data from approximately 1.5 million individuals to identify drug-diagnosis co-occurrences ( 736,000 pairs), significant associations (7,763 pairs), and assess direct drug usage through medical expert curation. The final resource comprises 7,763 associations with odds ratios > 3.5, manually annotated by six independent clinicians (3 in the UK and 3 in Denmark). Finally, the clinician annotations were scored using an Expectation-Maximization-based framework providing a confidence score for each pair. Our resource shows that the vast majority of significantly associated drugs and diagnoses in healthcare records are not due to direct treatment of the diagnosis. Additionally, through the annotation results, we demonstrate the importance of accounting for systematic differences among annotators when working with real-world data.

## 1 INTRODUCTION

Electronic health records (EHRs) contain decades of longitudinal clinical data that can enable large-scale biomedical research.[1, 2] However, healthcare systems differ across countries and evolve over time, creating heterogeneous EHR datasets with differences in coding practices and clinical context.[3, 4] Interpreting drug use across such data requires a standardized resource that captures real-world drug usage patterns across both primary and secondary healthcare systems. Terminology and classification systems such as ICD (International Classification of Diseases) are widely used in hospitals worldwide for diagnostic, administrative, and billing purposes.[3, 5] The ATC (Anatomical Therapeutic Chemical) system is another internationally adopted medication coding system used across clinical databases and EHRs.[6, 7] The combination of the ICD and ATC systems provides a good common terminology for the representation of drugs and diagnoses across datasets.

Publicly available resources that link drugs to disease and phenotype ontologies, such as SNOMED CT and the Human Phenotype Ontology (HPO), are valuable for research. However, they are not interchangeable with the terminologies historically used in EHR data in many countries.[8] Ontologies such as HPO represent concepts and their semantic relationships at high granularity, whereas clinical terminologies such as ICD are used for the organization and reporting of episodes in the clinic. Many legacy EHR systems remain anchored in ICD-based diagnostic coding because they were designed for clinical documentation rather than research, and phenotype ontologies do not map explicitly to ICD terminologies. The ICD-11 revision tries to mitigate the issue by mapping to SNOMED CT,[9] but it is not widely implemented yet. A drug-use resource based on ICD and ATC codes could therefore provide a practical framework for studying medication use in historical EHR data generated before the transition to newer terminology systems.

Clinical coding systems used in EHRs change over time. In 2018, the UK transitioned to SNOMED CT as a unified coding framework, replacing Read codes, using the Dictionary of Medicines and Devices (dm+d) to describe drugs, most often as Virtual Medicinal Products (VMPs). The dm+d terminology is incorporated within SNOMED CT, supporting a globally maintained ontology.[4] Such transitions in coding systems highlight the need for a resource that describes routine drug use in clinical settings.

A few examples of large-scale drug-disease datasets exist. PrimeKG is a graph-structured dataset that represents drugs using DrugBank identifiers, while diseases are encoded using MONDO and HPO ontology terms.[10] DrugBank[11] is a comprehensive drug information resource, whereas MONDO and HPO[12] provide standardized ontologies for diseases and phenotypic abnormalities. Although PrimeKG effectively links drugs to disease concepts, these ontology-based disease representations do not directly align with clinical terminologies. Consequently, integrating PrimeKG with real-world clinical datasets requires additional mapping between ontology terms and clinical terminologies, which can introduce errors and mapping ambiguity. ClinGraph is another graph-structured dataset that integrates biomedical concepts, including drugs and diseases.[8] However, ClinGraph does not provide explicit links between ATC drug classifications and ICD disease codes either. Instead, these entities are indirectly connected through PheCodes, which are phenotype groupings that aggregate related ICD codes into clinically meaningful disease categories commonly used in phenome-wide association studies.[13] While PheCodes facilitate large-scale phenotype analyses, they reduce coding granularity by collapsing multiple ICD codes into broader disease groupings.[13] As a result, relationships inferred through PheCodes may obscure specific drug–disease relationships. These limitations highlight the need for a contemporary disease-to-drug relationship map that reflects clinical care routines, enabling researchers to work with real-world healthcare data. Finally, MEDI[14] is a medication-indication resource that links drugs to their therapeutic indications. It is built from the public resources RxNorm, SIDER, MedlinePlus, and Wikipedia and represents drugs annotated by RxNorm concepts and diseases using UMLS and ICD-9 codes. Although this resource supports medication indication mapping, it does not include empirical observations of drug-diagnosis co-occurrences.

When working with real-world EHRs, it is important to distinguish between clinically directed drug use and non-directed drug use. Neither co-occurrence nor association between drugs and diagnoses can be interpreted as therapeutic indications. We distinguish between clinically directed drug use and indirect relationships, which are a result of treating comorbidities, prophylaxis, treatment of symptoms, or treatment of side effects from previous treatment. Our clinically curated resource will therefore allow us to distinguish between directed drug use and indirect relationships.

To the best of our knowledge, a comprehensive drug-use resource linking ATC drug codes to ICD diagnosis codes does not exist in the public domain. In this work, we generated a data-driven mapping of drugs (prescriptions and dispensations) to disease diagnoses in primary and secondary care. We used two large real-world healthcare datasets spanning different healthcare modalities and geographic regions (UK and Denmark) to generate a data-driven list of drug usage by disease using ATC and ICD-10 codes. Clinicians independently annotated the identified relationships, and scoring metrics measured their agreement while accounting for uncertainty. With this resource, we aim to support cross-disease and cross-country comparisons and provide a reference resource for translational research across multiple healthcare fields.

## 2 RESULTS

### 2.1 Cohort descriptions

For the Danish hospital data, we started with 2,551,705 patients and removed entries with missing ICD-10 and ATC codes, as well as ICD-10 codes describing events, external causes, and healthcare utilization. The remaining 1,327,277 patients gave rise to 478,227 drug-diagnosis pairs [Table 1]. Those pairs were filtered for prevalence in 50 unique patients (58,246 pairs) and then tested for significant association using Fisher’s exact test. Afterward, multiple test correction was applied using the False Discovery Rate - Benjamini-Hochberg (FDR-BH) approach, leaving 6,434 (11.04%) associations remaining under the 0.05 significance level and 3.5 odds ratio thresholds.

For the UK general practitioner (GP) data, the initial population was filtered in the same way, from 230,523 to 225,522 participants with 258,237 drug-diagnosis pairs. After the prevalence filter, 18,046 pairs remained; of those, we identified 1,615 (8.95

### 2.2 Distribution of ATC and ICD-10 chapters in primary and secondary care

Overall, the UK and DK data showed distinct differences in the distribution of both ICD-10 diagnoses and ATC drug codes represented in the significant drug-diagnosis pairs [Figure 1c and 1d]. It should be noted that the UK data is derived from primary healthcare data, whereas the DK data comes from a secondary healthcare (hospital) setting. Within the significant associations between ICD-10 and ATC codes, the DK data showed the highest percentage of ICD chapter II, neoplasms (DK = 22.32%, UK=1.13%). This is consistent with the source of the DK data being hospital records, where treatment of cancers and neoplasms will be common. In contrast, the UK data showed the highest percentages for ICD-10 chapter I, certain infectious and parasitic diseases (UK=10.12%, DK=4.74%), chapter V, mental and behavioral disorders (UK=9.69%, DK=2.70%), and chapter X, diseases of the respiratory system (UK=11.12%, DK=7.65%).

**Figure 1:**
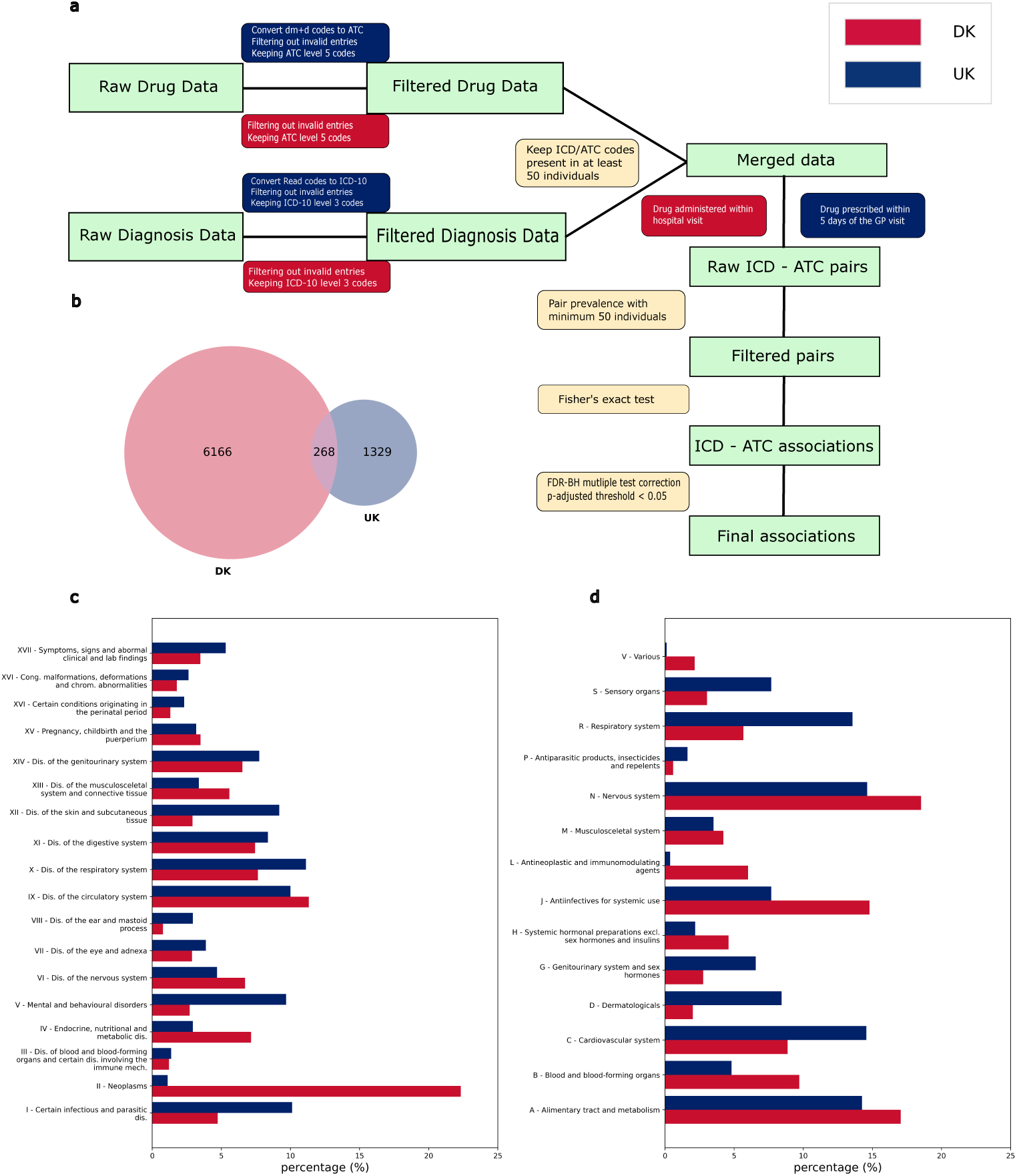
Data and analysis overview. a) The analysis plan implemented was the same for both UK and DK datasets. Due to small differences in the input data, some steps performed additional code mapping or processed the data differently for the same outcome. These differences are denoted by the different colors on either side of the schematic, with blue for the UK and red for DK. Overall, the same thresholds and filtering were applied to both. b) Overlap of significant pairs identified in the two datasets. c) ICD-10 chapter prevalence in the final UK (blue) vs DK (red) pairs. d) ATC chapter prevalence in the final UK (blue) vs DK (red) pairs.

Within significant associations between ICD-10 and ATC codes, the DK data had the highest percentage of ATC chapter N, nervous system (DK=18.53%, UK=14.64%) [Figures 1c and 1d]. This likely reflects the inclusion of anesthetics and analgesics in this chapter, which are prevalent in hospital prescriptions.[15] The UK data had a higher percentage of ATC chapters C, cardiovascular system (UK=14.58%, DK=8.86%), and R, respiratory system (UK=13.56%, DK=5.6%). This is likely to reflect the treatment of chronic diseases in a primary healthcare setting, including arrhythmia, angina, and coronary artery disease, as well as asthma and chronic obstructive pulmonary disease. Notably, ICD-10 chapter C, neoplasms, and ATC chapter L, antineoplastic and immunomodulating agents, are almost absent from the UK data, consistent with the lack of use of these drugs in primary care.

### 2.3 Clinician annotations and confidence scores

The significantly associated drug-diagnosis pairs identified in each country were reviewed independently by three clinicians with experience in the respective country’s healthcare system: three DK clinicians reviewed the DK pairs and three UK clinicians reviewed the UK pairs. Inter-annotator agreement was assessed separately for the Danish and UK datasets using Fleiss’ kappa. The DK clinicians showed moderate agreement, with a kappa of 0.545. The UK clinicians achieved a kappa value of 0.283, interpreted as fair agreement.[16, 17]

In the Danish annotations, complete agreement was most commonly observed for pairs classified as not directly prescribed for the diagnosis [Figure 2a]. All three clinicians assigned a “no” label to approximately 65% of reviewed pairs (3,974 pairs). This implies the majority of disease-drug pairs are straightforward cases where drug usage is not directly related to the disease specified. Complete agreement on direct prescriptions was less frequent, with all three clinicians assigning a “yes” label to 10.78% (694 pairs). This likely reflects both the smaller number of associated pairs that reflect true direct drug usage and clinician conservativeness. Comparatively, the most frequent complete consensus pattern in UK data was agreement on “yes” annotations, observed for 20.84% (333 pairs) [Figure 2b]. This might be influenced by the smaller size of the UK data but also suggests that UK clinicians were more inclined toward positive relationship assignments than DK clinicians. This may reflect the source of the drug-disease pairs - in DK from a hospital setting, in the UK from a primary care setting - and the specificity of treatment that occurs in each environment.

**Figure 2:**
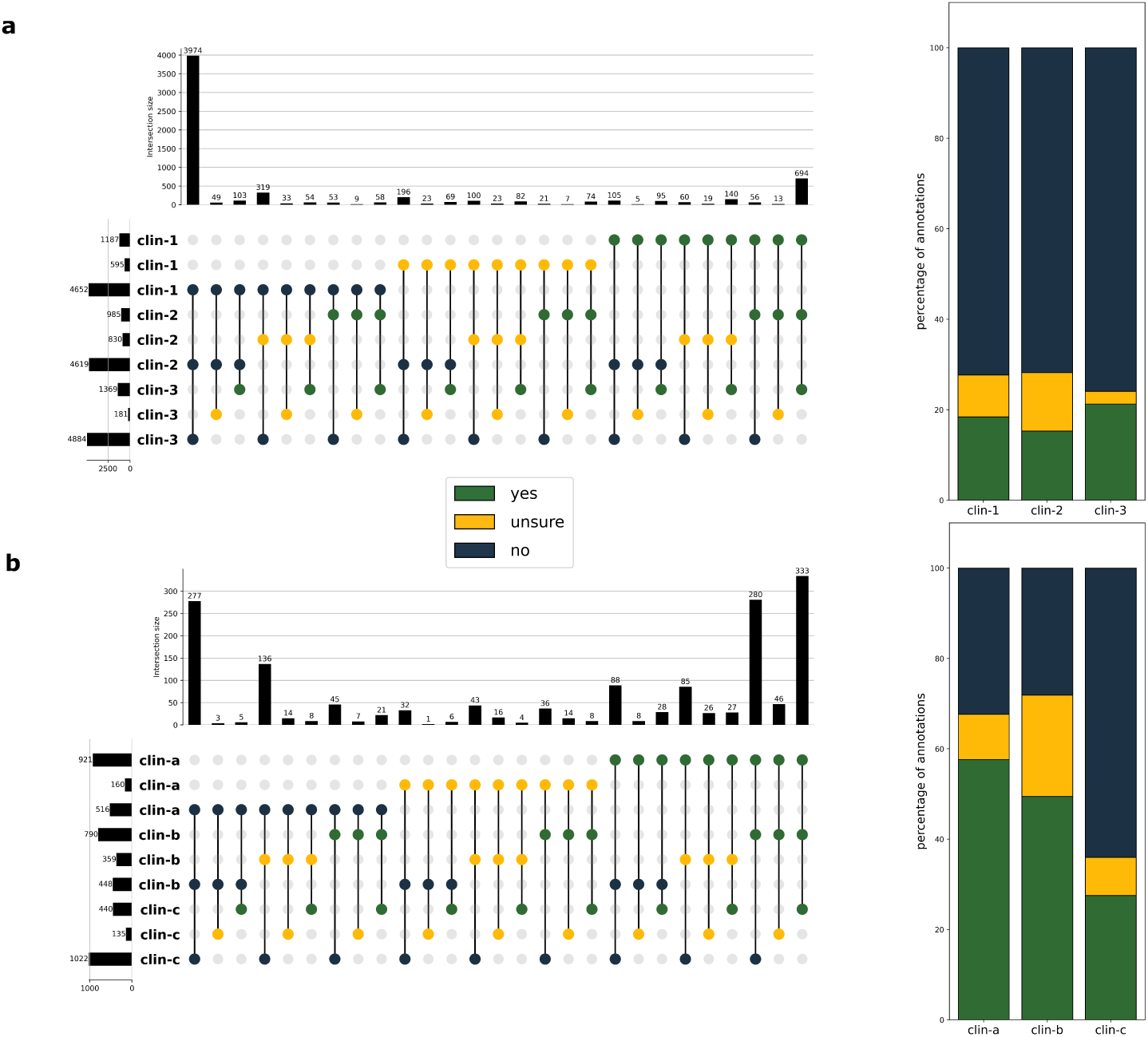
Clinician overlap in annotations and clinician annotation preferences (“yes” - green, “unsure” - yellow, “no” - blue). a) Danish clinician UpSet plot with three rows per clinician based on the annotation options (left) and barplot of the annotation percentages per clinician (right). b) UK clinician overlap (left) and preference percentages (right).

Within each country set, clinician-specific patterns were observed, potentially reflecting differences in the conservativeness of label annotation and ambiguity of drug usage for specific diseases. In the DK data, the “unsure” label was used most frequently by clinician 2, who assigned this label to 830 pairs. In 55% (459 pairs) of these cases, the other two clinicians both agreed upon a “no” or “yes” annotation. This suggests that clinician 2 was the most conservative of the three clinicians in assigning definite associations. In the UK data, patterns were more complex, reflected in the lower Fleiss’ kappa. UK clinicians A and B assigned “yes” labels to more than half of the reviewed pairs, whereas clinician C predominantly assigned “no” labels. The next most common pattern involved “yes” labels from clinicians A and B and a “no” label from clinician C, observed for 280 pairs. This indicates that clinicians A and B have a common prior that is different from that of clinician C.

The signed confidence score was calculated for each drug-diagnosis pair within each country using the Dawid-Skene approach (see methods). The distribution [Supp fig: confidence score distributions] suggests that Danish pairs had a higher density of negative scores, whereas UK pairs had a higher density of positive scores. This observation aligns with the “yes” and “no” percentages UK vs DK clinicians assigned to the individual pairs. In addition, scores for UK pairs showed a higher density around zero, particularly between -0.5 and 0.5, than for DK pairs. Higher density around zero suggests greater overall uncertainty in pairwise annotations among UK clinicians.

### 2.4 Overview of ICD and ATC chapter combinations

In the DK associations, multiple ICD-ATC chapter combinations had a high absolute negative average score, meaning that the dataset contained fewer pairs of directed drug use. These include ICD chapter II (Neoplasms) and ATC chapter A (Alimentary tract and metabolism), chapter II and chapter J (Antiinfectives for systemic use), and chapter XIII (Diseases of the musculoskeletal system and connective tissue) and chapter N (Nervous system). These are chapter pairs containing diseases that appear with comorbidities; for example, insulin (A10AB05) use for pancreatic cancer (C25) strongly indicates a diabetes comorbidity (E11), or use of oral vancomycin (A07AA09) in hematological malignancies (C90 and C91) reflects that patients suffer from Clostridium difficile (A04) in the intestines, which is a common complication of chemotherapy-associated immunosuppresion rather than treatment of the malignancy itself. In these cases, the drugs targeting the comorbidity chapter have a high frequency and lead to high absolute negative average scores. In contrast, we also observe combinations with lower counts of overlap among ICD-10 chapters and ATC chapters that have very high positive average scores. Such cases include ICD-10 chapter XII and ATC chapter D, chapter XIII and chapter H, and chapter III and chapter B. These indicate ICD-10 chapters with diagnoses that often appear in isolation and are treated with a small subset of highly targeted drugs.

Across ICD-ATC chapter combinations in the UK data, most combinations contained relatively few significant drug-diagnosis pairs; however, the combinations show high average confidence scores across both frequent and infrequent combinations [Figure 3b]. The largest high-confidence associations were found for clinically coherent chapter pairs, including ICD chapter VI (Diseases of the nervous system) with ATC chapter N (Nervous system) (54 pairs, 3.38%), and chapter IX (Diseases of the circulatory system) with ATC chapter C (Cardiovascular system) (111 pairs, 6.94%). This provides a good positive control. Some high-count combinations showed comparatively lower average confidence scores, including chapter V (Mental and behavioral disorders) with chapter N (Nervous system), and chapter X (Diseases of the respiratory system) with chapter R (Respiratory system). This could reflect less standardized care for these complex conditions, lesser-known etiologies, and is likely also a reflection of the lower Fleiss’ kappa amongst the UK clinicians. Relative to the Danish data [Figure 3a], the UK data covered fewer ICD–ATC chapter combinations and contributed fewer significant pairs within each combination. This is likely due to the smaller size of the UK primary care data compared to the DK hospital data.

**Figure 3:**
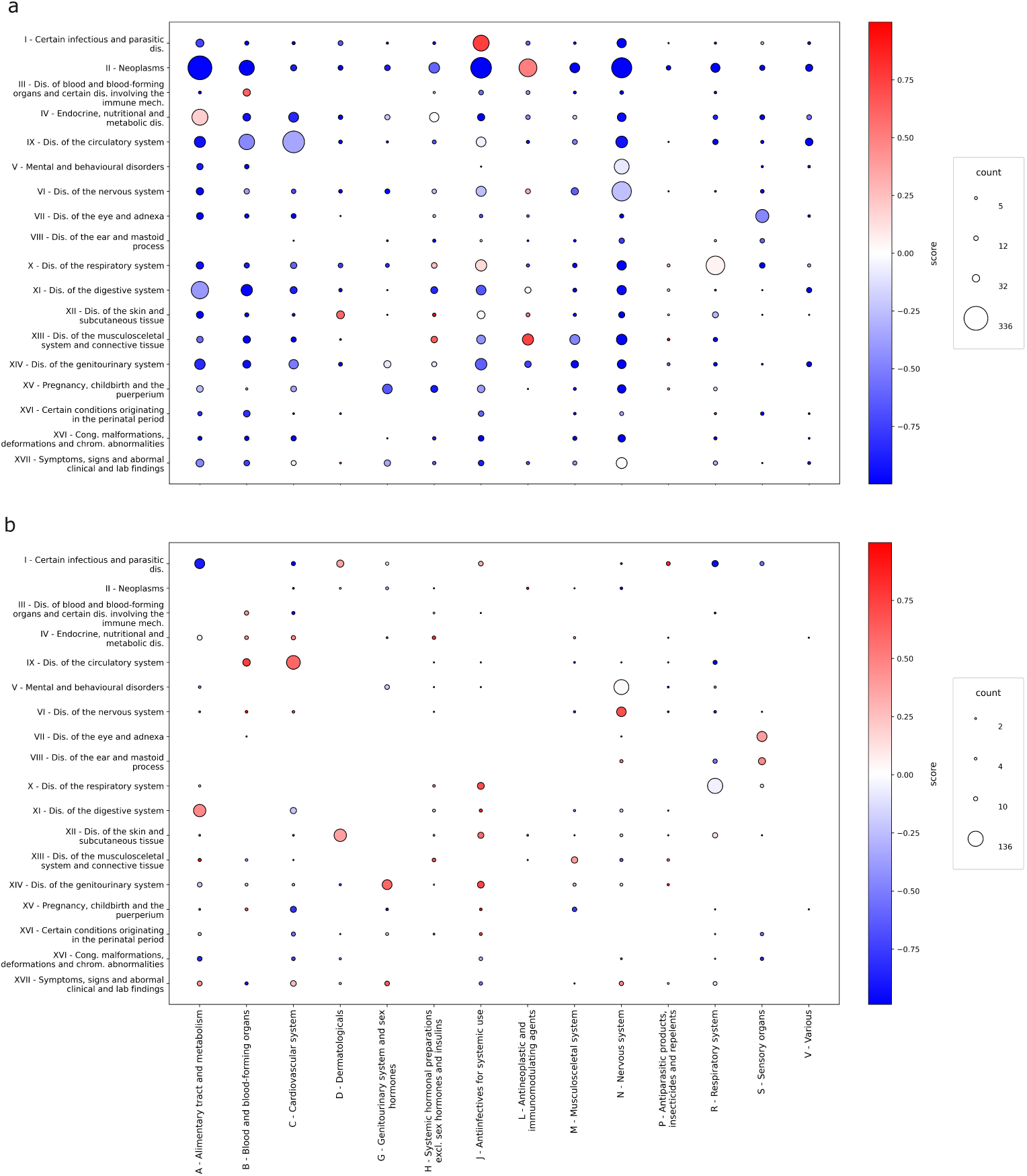
Average score and number of pairs per ICD-10 and ATC chapter combination. The size of each dot indicates the number of ICD-ATC pairs that belong to that chapter combination. The color map indicates the average confidence score for pairs in that chapter combination. More intense colors indicate stronger confidence, with red colors indicating a higher average confidence towards “yes” annotations, while lower average confidence scores are colored blue. a) Significant pairs from the Danish dataset. b) Significant pairs from the UK dataset.

### 2.5 Pair overlap between primary and secondary care

To assess the clinical similarity of the data, we looked into the intersection of the two datasets. There is a small overlap of 268 pairs (4.16% of the total DK pairs, and 16.78% of the total UK pairs) common across the two datasets [Figure 1b]. Of the significantly associated drug-diagnosis pairs across both datasets, there were more pairs that all six clinicians (three from DK, three from UK) agreed on a “yes” label than a “no” label. Additionally, there were no pairs that all clinicians labeled “unsure”. This indicates that pairs that are present in both datasets are more likely to represent direct drug usage pairs. However, the agreement across all six clinicians on those pairs was assessed as weak, with a Fleiss’ kappa value of 0.273. This is likely due to the fact that there are a lot of annotators, and though most of them often agree, it is a lot harder to have overall agreement amongst them.

The most prevalent ICD-10 and ATC chapter combinations in the common pairs among the two countries were ICD chapter IX (Diseases of the circulatory system) with ATC chapter C (Cardiovascular system) (38 pairs, 14.18%), chapter X (Diseases of the respiratory system) with chapter R (Respiratory system) (27 pairs, 10.07%), and chapter XI (Diseases of the nervous system) and chapter N (Nervous system) (22 pairs, 8.21%). These findings suggest that drug–diagnosis pairs shared across countries and healthcare systems predominantly reflect clinically concordant ICD and ATC categories.

## 3 DISCUSSION

### 3.1 Annotated drug-use map

In this study, we mined co-occurrence associations between diagnoses and drug information from real-world healthcare data, spanning primary care and hospital admissions in two different healthcare systems. In total we identified 7,763 unique, significantly associated drug-diagnosis pairs with high effect sizes and performed expert annotation of these pairs for direct treatment. To enable easy integration of the resource with data-driven and machine learning pipelines, the annotations were also combined into a single signed confidence score. The resulting drug-use map is a data-driven, curated dataset that may be used as a resource for large-scale EHR research.

### 3.2 The majority of drug-diagnosis associations are not due to direct drug usages

The expert annotations show that statistical association alone is not equivalent to direct drug use. Only a small fraction of the significantly co-occurring pairs in the final dataset were agreed upon by all experts as cases of direct treatment, but more than half of the pairs were agreed upon by all as non-direct treatment. Some pairs are likely attributable to symptomatic treatment; for example, baclofen (M03BX01) relieves spasticity associated with multiple sclerosis (G35) without treating the underlying disease. Others are indicative of prophylactic use; for example, topiramate (N03AX11) can be used as a prophylactic for migraine. Some will further represent treatment of side effects caused by other medications (especially in cancer), or even treatment of comorbid diseases, for example, folic acid (B03BB01), which is often prescribed with methotrexate (L04AX03) to reduce its toxicity in rheumatoid arthritis (M05) patients; enofibrate (C10AB05) prescribed for co-occurring dyslipidemia (E78) in patients with hypertension (I10); or ondansetron (A04AA01) prescribed to manage chemotherapy-induced nausea and vomiting in patients with pancreatic cancer (C25).

### 3.3 Reviewing different healthcare system modalities across two countries

The identified pairs originate from two distinct healthcare data modalities, with marked differences in dataset scale, clinical setting, and coding patterns. The Danish dataset, based on hospital records, contributed 82.8% of the significant drug–diagnosis pairs, likely reflecting its larger patient population and more detailed documentation of encounters. Meanwhile, 17.2% of the results were identified in the UK dataset, which was derived from primary care records.

We identified clear differences between the ICD-10 and ATC codes found in the two original datasets. The Danish diagnosis codes consisted mostly of conditions mainly treated in a hospital setting, including Neoplasms (II), Circulatory system diseases (IX), and Endocrine, nutritional, and metabolic diseases (IV). In contrast, the UK data were enriched for conditions commonly assessed and managed in routine primary care, such as Certain infections and parasitic diseases (chapter I) and Respiratory system diseases (X). As expected, the ICD chapter II (Neoplasms) was almost exclusively present in the hospital data and almost completely absent from the GPs. Additionally, neoplasms had many significantly associated drugs, but only antineoplastic drugs received a high confidence score. Treatment of neoplasms often involves multiple side effects, resulting in patients receiving drugs to manage those. These associations received negative scores, indicating non-direct treatment of the index condition.

Furthermore, the differences between the two datasets extend to the drugs identified in them. The hospital data from DK showed a high frequency of Nervous system drugs (N) and Anti-infective drugs (J). The most frequent nervous system drugs are analgesics and anesthetics, which are used in surgeries and/or to manage pain afterward. Additionally, generic anti-infective drugs are often used to prevent infections either proactively or after the fact in a hospital setting. As a result, nervous system and anti-infective drugs appear in multiple pairs across all diagnosis chapters and therefore received low scores, indicating that most use-cases are preventative or aimed at alleviating symptoms such as pain.

By including data from both countries, we are able to generate a resource that covers a wider variety of conditions and treatment scenarios as well as more cases of frequent non-directed drug usage. Although the differences in both geographical settings and the scope of the healthcare systems render the two datasets non-comparable, their integration broadens the resulting resource’s inclusion of clinical treatment patterns. Additionally, primary and secondary EHRs are often treated as different modalities, which hinders the applicability of the results in a holistic setting. Thus, our work provides a resource for analyses of both primary and secondary EHRs by including information from both. An ideal expansion of this work would be to add primary healthcare data from Denmark and hospital diagnoses and drug administrations from the UK, both for replication purposes and to expand our final list. However, this is not possible within the scope of the current study.

### 3.4 Observing consensus among experts

When quantifying consensus among our clinician annotations between drugs and diagnoses, we found moderate agreement. This serves as an indicator that real-world applications of drug usage are often context-dependent and drug labels do not suffice. Furthermore, it shows that most drugs are not used to treat a specific disease, but to treat symptoms of a disease, comorbidities occurring alongside a disease, side effects of the specific treatment, or for prophylactic reasons. Finally, the moderate level of agreement suggests that annotations were influenced by differences in clinicians’ experience and interpretation. Incorporating assessments from multiple clinicians and explicitly accounting for inter-rater variability is therefore important for modeling an underlying prior of the “truth”.

More importantly, it highlights the need for curated ground-truth datasets generated from real-world observations that reflect actual drug use. In particular, the high level of disagreement among multiple experts highlights the nuances involved in creating reference datasets and the need for greater clinical granularity. Rather than forcing consensus, the Dawid-Skene EM approach allowed us to score the pairs based on the annotations while incorporating clinician-specific patterns and overall agreement. In this way, we were able to model annotator preferences, and thereby lessen the impact of highly divergent annotation patterns.

### 3.5 Comparison with preexisting resources

Our resource complements existing drug-use resources by shifting focus from literature-based approaches that use data such as drug indications to real-world data-driven approaches, i.e., drug-diagnosis co-occurrences. It could therefore be combined with a resource like MEDI[14], which integrates information across literature. Such integration would enable the discovery of patterns aside from direct drug use, like symptom treatment or treatment of comorbidities, along with literature validation.

Our annotation strategy also differs from previous MEDI efforts in both scale and structure. Whereas MEDI’s pairs were curated by two clinicians, our study incorporated pair-level assessments from six clinicians and applied a probabilistic label-aggregation model to account for annotator-specific tendencies and disagreement. As a result, the signed confidence score provides not only a summary estimate of clinical interpretability but also represents pair-level ambiguity, which should always be considered when dealing with heterogeneous real-world data for which the underlying truth is unknown.

Our study also extends literature-based mapping approaches, such as that of Choi et al.[5], by identifying and annotating real-world ICD–ATC associations across two healthcare systems rather than relying solely on previously documented medication–indication links. This distinction is important because EHR-derived associations reflect how medications are used within specific clinical settings, including differences between primary and secondary care. By combining empirical co-occurrence patterns with expert annotation, the resulting map provides a more granular representation of drug–diagnosis relationships and their interpretability in routine clinical data, which is a resource that has been lacking in large-scale EHR research.

Beyond distinguishing direct treatment from associations-by-proxy, our resource provides a more granular representation of the clinical complexity underlying drug-diagnosis pairs. The signed confidence score may support the development and evaluation of clinical predictive models by serving as a continuous target, allowing models to learn from both directed treatment pairs as positives and associations-by-proxy (negatives), which can be especially beneficial when studying comorbidity-related prescribing. These well-documented positives and negatives are particularly important in the age of large language models (LLMs), massive knowledge graphs, and retrieval-augmented generation (RAG) algorithms, which contain large amounts of information of varying degrees of confidence. In addition, the inclusion of data from distinct healthcare systems and countries captures setting-specific variation in prescribing and coding practices, supporting the evaluation of model generalizability across clinical contexts.

## 4 FUTURE PERSPECTIVES

Our work focuses on generating a drug use map for commonly observed drug-diagnosis co-occurrences. Thus, we use highly prevalent drug and diagnosis codes by excluding both codes and pairs below a specific patient count, as they may be susceptible to noise and are less generalizable overall. Future work could investigate rarer drug-diagnosis pairs to extend the scaffolding established in this study. Drugs recorded more sparsely in healthcare records may indicate more directed prescription patterns and should be present in an extensive drug-use resource. Additionally, given the substantial clinical annotation burden, we prioritized associations with larger effect sizes as a subgroup, but clinically meaningful pairs with lower effect sizes can contain drugs that are used exclusively in specific patient subgroups and are therefore not able to show higher effect sizes. Building on our work, future studies could utilize LLMs and GraphRAG algorithms to prioritize drug-diagnosis pairs for review or involve more clinical experts in the annotation process, which would also offer more insight into clinician agreement. Although LLMs are now frequently used in research, they remain unreliable and can omit important associations. Conversely, adding more clinician annotators could increase the size of the curated dataset but would require substantial personnel time. Future work could validate and expand our resource by incorporating additional datasets from different countries. Ideally, such efforts would include primary and secondary healthcare data from the same country, allowing for an additional comparison between healthcare system settings. Such a study would further describe the differences in drug use patterns among the two systems without confounders such as country-specific differences in coding practice.

## 5 CONCLUSION

In conclusion, this study has identified 7,763 significantly associated drug-diagnosis pairs in EHR data. These drug-diagnosis pairs were annotated to support large-scale analyses on real-world data. By integrating annotations from six clinical experts, the signed confidence score provides a pairwise measure of clinical interpretability that accounts for clinician-specific annotation tendencies and inter-rater disagreement. The observed differences between the UK primary care and Danish secondary and tertiary care datasets were consistent with expected variation between general practice-recorded and hospital-based drug use, underscoring the importance of healthcare context when interpreting EHR-derived associations. This resource provides an empirically grounded reference set for the development, evaluation, and validation of data-driven models using large-scale EHR data.

## 6 MATERIALS AND METHODS

### 6.1 Study population

The Danish data include diagnoses from the Danish National Patient Registry (DNPR) and prescriptions from 12 public hospitals across Eastern Denmark (the Capital Region of Denmark and Region Zealand).[6] DNPR comprises diagnoses for each admission to a Danish hospital, as ICD-10 codes, which are merged with an EHR dataset of in-hospital-administered prescription data, as ATC codes. Diagnosis data were available for approximately 2.5 million individuals, and prescription data for 1.3 million patients from 2008 to 2016. Analysis was restricted to patients present in both sources, yielding a dataset of 1.3 million patients over the period 2008-2016. The diagnoses in DNPR are timestamped by hospitalization periods with admission and discharge dates, along with the assigned episode diagnoses. The medication data were recorded as discrete drug-administration events, providing greater confidence that the recorded medications were administered to patients rather than merely prescribed.

UK Biobank (UKBB) is a large-scale cohort of approximately 500,000 participants recruited from across the UK.[18] Our analysis included approximately 225,000 individuals who had consented to EHR linkage. We used diagnosis and prescription data from GPs, with diagnoses recorded in Read version 2 codes and prescriptions encoded in dm+d, Read version 2, or British National Formulary (BNF).

### 6.2 UK code conversion to ICD-10 and ATC terms

Read codes are a hierarchical clinical coding system used by GPs in the UK to record diagnoses and symptoms (e.g., G20..00 for essential hypertension). Prescription records were standardized to dm+d VMP codes following the procedure in Ytsma et al.[19, 20] for 72% of records (n=41,238,074). VMP codes represent the medicinal product independent of brand (e.g., 42109611000001109 for 500 mg paracetamol tablets). Pair identification was restricted to 1964-2017 because the data were sparse prior to 1964 and unavailable after 2017.[20]

The UKBB data contained Read and VMP codes which were mapped to ICD-10 and ATC codes, respectively, using official NHS mappings. The Read-to-ICD-10 mapping was based on the appropriate file (NHS-TRUD “Coding system lookups and mappings version 4 (June 2023)”), and the VMP-to-ATC mapping was based on the appropriate corresponding file (2024_7.2.0/f_bnf1_0110724.xml), matching the corresponding input data versions. After mapping, records with missing patient IDs, dates, diagnoses, or prescription codes were excluded from downstream analyses, leaving 225,522 individuals for downstream analysis.

### 6.3 Data preparation and pair identification

From both datasets, we removed ICD-10 codes in chapters XIX, XX, XXI, and XXII from the diagnosis registry, as in previous studies[21], because they describe external events, circumstances, and healthcare interactions rather than disease entities and offer little diagnostic value, while increasing complexity and dimensionality [Figure 1a].

In the UKBB data, ICD-10 diagnosis codes and ATC prescription codes were mapped and then merged using participant identifiers. Diagnosis-prescription pairs were defined as co-occurring when the prescription was recorded within five days of the diagnosis date. The five-day distance window was chosen to balance clinical specificity with practical prescribing workflows, capturing prescriptions relative to the recorded diagnosis while allowing sufficient time for further assessment by the GP.

### 6.4 Pair filtering and association testing

Prior to association testing, ICD-ATC pairs observed in at least 50 patients/participants within each dataset independently were included in downstream analysis. The prevalence size ensures that we focus on common pairs, supporting statistical disclosure control by avoiding very small patient groups and reducing the risk of spurious associations caused by sparse observations of either the diagnosis or the prescription [Figure 1a].

We performed Fisher’s exact testing across all diagnosis-prescription pairs. We tested the co-occurrence pairs independently for association and then corrected the resulting p-values using the false discovery rate Benjamini-Hochberg (FDR-BH) approach. To limit our resource to significant associations with strong effect sizes, the association-tested pairs were filtered for corrected p-value < 0.05 and odds ratio > 3.5.

### 6.5 Relationship annotation

A given drug may be prescribed for several diagnoses, and a given diagnosis may be treated by multiple drugs. Therefore, significant drug–diagnosis pairs identified in each dataset were reviewed by country-specific clinicians to assess whether the observed associations reflected direct treatment of the indexed diagnosis. In total, six clinicians participated in the annotation process: three DK clinicians reviewed pairs identified in the DK dataset, and three UK clinicians reviewed pairs identified in the UK dataset.

Each clinician independently annotated the pairs from their respective country as “yes,” “no,” or “unsure.” Pairs were labeled “yes” when the drug was prescribed specifically to treat the indexed diagnosis, “no” when the association likely reflected treatment of a comorbidity, symptom, or downstream condition, and “unsure” when the clinician could not confidently classify the relationship. After independent review, we derived a country-specific consensus annotation. We used this process to distinguish direct therapeutic associations from proxy associations in the final results.

### 6.6 Statistical analysis of annotations

We assessed the agreement among clinician annotations using Fleiss’ kappa coefficient, which quantifies the inter-annotator agreement among more than two annotators. Fleiss’ kappa measures the extent to which clinicians agree on labeling pairs, relative to what would be expected by chance. The resulting kappa statistic was interpreted according to the previously established ranges by Landis and Koch.[16, 22]

To account for clinician disagreement, we defined a scoring system that reflects the magnitude and directionality of annotation consensus for each drug-diagnosis pair. Clinician annotations were aggregated into a single pair-level score using a probabilistic label-aggregation framework based on the Dawid–Skene model.[23] In this framework, the true class of each pair is treated as a latent variable while the observed annotations are modeled as noisy realizations of the latent class. The model estimates both the posterior probability of each latent class for every pair and the clinician-specific labeling parameters. Thus, compared to simple majority voting, the Dawid-Skene model explicitly learns clinician-specific labeling patterns. Each clinician gets a labeling profile per class (“yes”/“unsure”/“no”), which corresponds to the probability of assigning a given label conditional on the probability of the latent class. This allows the aggregation procedure to distinguish between pair-level uncertainty and clinician-specific labeling patterns.

Model parameters were estimated using the Expectation-Maximization algorithm. In the expectation step, the posterior probability that each pair belonged to the direct treatment or association-by-proxy class was computed, conditional on the observed annotations and the current estimates of clinician labeling patterns. In the maximization step, these posteriors were used to update both the class prevalence and the clinician-specific annotation matrices. These steps were repeated iteratively until convergence, with the final posterior probabilities serving as the scores.

The final posterior probabilities were transformed into a signed continuous confidence score that ranged from -1 to +1, with higher absolute values indicating greater confidence (magnitude) and the sign indicating the directionality (negative for associations-by-proxy and positive for direct treatment; Table 1).

## ACKNOWLEDGMENTS

Acknowledge contributors who do not meet authorship criteria.

## DATA AVAILABILITY

The data underlying this article will be made available upon publication.

